# Evaluation of lockdown impact on SARS-CoV-2 dynamics through viral genome quantification in Paris wastewaters

**DOI:** 10.1101/2020.04.12.20062679

**Authors:** S Wurtzer, V Marechal, JM Mouchel, Y Maday, R Teyssou, E Richard, JL Almayrac, L Moulin

## Abstract

SARS-CoV-2 is the etiological agent of COVID-19. Most of SARS-CoV-2 carriers are assumed to exhibit no or mild non-specific symptoms. Thus, they may contribute to the rapid and mostly silent circulation of the virus among humans. Since SARS-CoV-2 can be detected in stool samples it has recently been proposed to monitor SARS-CoV-2 in wastewaters (WW) as a complementary tool to investigate virus circulation in human populations. In the present work we assumed that the quantification of SARS-CoV-2 genomes in wastewaters should correlate with the number of symptomatic or non-symptomatic carriers. To test this hypothesis, we performed a time-course quantitative analysis of SARS-CoV-2 by RT-qPCR in raw wastewater samples collected from several major wastewater treatment plant (WWTP) of the Parisian area. The study was conducted from 5 March to 23 April 2020, therefore including the lockdown period in France (since 17 March 2020). We confirmed that the increase of genome units in raw wastewaters accurately followed the increase of human COVID-19 cases observed at the regional level. Of note, the viral genomes could be detected before the beginning of the exponential growth of the epidemic. As importantly, a marked decrease in the quantities of genomes units was observed concomitantly with the reduction in the number of new COVID-19 cases which was an expected consequence of the lockdown. A s a conclusion, this work suggests that a quantitative monitoring of SARS-CoV-2 genomes in wastewaters should bring important and additional information for an improved survey of SARS-CoV-2 circulation at the local or regional scale.

## Introduction

SARS-Cov-2 is a positive-sense single-stranded RNA virus of the *Coronaviridae* family and the etiologic agent of COVID-19, a globalized infection affecting more than 2.5 million people worldwide and causing more than 180 000 deaths in total, including more than 160 000 cases in France on April 22, 2020. Virus transmission is mainly associated with the projection of respiratory droplets although a possible contamination through aerosols, contaminated hands and inert surfaces is likely. SARS-Cov-2 causes severe complications mostly in elderly or people suffering from comorbidity factors (such as diabetes, hypertension, obesity, acquired or iatrogenic immunosuppression).

The viral infection may initiate in the upper respiratory and/or the lower respiratory tracts. Similarly to SARS-CoV-1 (1) and MERS-CoV (2), SARS-CoV-2 genome was also detected in blood and stools (3–5). This argue for a possible enteric phase of the infection although isolation of infectious virus from feces seems difficult (6). Of note diarrhea have been reported in some cases of COVID-19 (6). Importantly, SARS-CoV-2 genomes could be detected in feces several weeks after it could not be detected anymore in oral swabs, suggesting that viral excretion in tools may be longer than oral secretion (7). The presence of viral genomes in stools may open new perspective in the survey of SARS-CoV-2 carriers. It would notably suggest that the virus could possibly be transmitted by a feco-oral route, an hypothesis that should likely deserve a careful examination (8).

Management of an epidemic, such as lockdown decision, requires a careful monitoring of the infected population by detecting the virus in carriers through massive or targeted testing. Investigating the proportion of people that have been infected through sero-epidemiological surveys is equally important but antibodies against SARS-CoV-2 will appear only weeks after initial infection (9,10). In the case of COVID-19, due to the lack of systematic and repeated screening of the population, the precise number of infected people is difficult to assess, especially because of the high proportion of infected people that exhibit only few or no symptoms but could secrete and silently transmit the virus (11–14). Depending on screening kit availability and public health policy, testing strategy varies between countries which may explain some discrepancy between worldwide data. Estimating the effective proportion of infected individuals is essential for monitoring the epidemic spread and to propose adapted and efficient control procedures, such as partial or total lockdown. France went into lockdown on the March 17 2020, a decision that was expected to have a major impact on virus circulation especially when asymptomatic carriers are considered to have a strong impact on virus transmission. This decision was motivated by the urgent need to limit exposure of so-called fragile people who are at highest risk to develop the most severe forms of the disease (11,13,15).

Analysis of raw wastewaters collected at the inlet of wastewater treatment plants (WWTP) may provide essential information on the health of the human population that is connected to the WWTP. It may notably allow measurement and identification of pathogens or drugs that may be difficult to assess otherwise. Using this method, the European Monitoring Center for Drugs and Drug Addiction follows drugs and their metabolites in the wastewater of several European cities (16). In addition previous works on human enteric viruses in urban river and in raw and treated wastewaters demonstrated that the presence of these viruses was directly linked to epidemic state in the population (17,18). This strongly argue for a close monitoring of fecal viruses in wastewater as a new and complementary tool for investigating human epidemics.

Enveloped viruses like coronaviruses are expected to be less resistant than naked viruses that are usually tracked in waste and environmental waters. There is still little information on the persistence of coronaviruses in waters and most of our current knowledge has been inferred from experiments made on surrogate viruses. Recent data suggested that infectious SARS-CoV-2 is particularly resistant in environmental conditions: 3.5 half-life days in the air, 7 days on some surfaces, any reduction at pH between 3 and 10 (6,19). Previous studies on SARS-CoV-1 indicated a significant persistence at 4 °C even in wastewater (more than 20 days), or a persistence of at least 1 or 2 days at summer temperatures (1,20,21).

Altogether these results led us and others to suggest that the detection of SARS-CoV-2 genomes in wastewater could provide an early and global tool to monitor virus circulation in addition to human epidemiological data (22–24).

A first publication underlined the putative benefit of a qualitative approach for monitoring SARS-CoV-2 in wastewaters (24). Other studies used quantitative measurements of viral genomes but the survey only started at the apex of the epidemic (25). Here, we used a specific reverse-transcription quantitative PCR (RT-qPCR) method to precisely quantify SARS-CoV-2 genome equivalents in raw wastewaters of the Parisian area. A 2-months survey covering the lockdown period allowed to observe an increase and a decrease in the total quantities of viral genomes that paralleled the number of new COVID-19 cases in the same region. To our knowledge, this is the first real-time indirect survey of SARS-CoV-2 circulation during a lockdown period.

## Methods

### Sample collection

Three WWTP (more than 100 000 inhabitants linked to the station from the Parisian area were sampled since the start of the epidemic (March 5^th^, 2020). Samples were kept at 4°C and processed less than 24 hours after sampling.

### Concentration methods

Samples were homogenized, then 11 ml were centrifugated at 200 000 × g for 1 hour at +4°C. Viral pellets were resuspended in 400 μL of PBS 1X buffer.

200µL of viral concentrate were lysed and extracted using PowerFecal Pro kit (QIAGEN) on a QIAsymphony automated extractor (QIAGEN) according to a modified manufacturer’s protocol. Extracted nucleic acids were filtered through OneStep PCR inhibitor removal kit (Zymoresearch) according the manufacturer’s instructions.

### Molecular detection method

The RT-qPCR primers and PCR conditions used herein have been previously described (26). The amplification was done using Fast virus 1-step Master mix 4x (Lifetechnologies) with oligonucleotide concentrations recommended previously. Detection and quantification were carried on the E gene by RT-qPCR. Positive results were confirmed by amplification of a region located within the gene encoding for the viral RNA-dependent RNA polymerase (RdRp). An internal positive control (IPC) was added to evaluate the presence of residual inhibitors. The detection limit was estimated to be around 10^3^ genome units per liter of raw wastewater.

The quantification was performed using a standard curve based on synthetic oligonucleotide corresponding to the full-length amplicon on the E gene (SARS-Cov2 Wuhan-Hu isolate sequence NC_045512.2). Amplification reaction and fluorescence detection were performed on Viaa7 Real Time PCR system (Lifetechnologies).

### Modelization of Viral RNA excretion

Even if the real number of infected people is unknown, we attempted to compute the expected amount of viral RNA shed in stools, using available information. Since the very beginning of the epidemic, the daily number of patients consulting at the emergency department of greater Paris hospitals and diagnosed with COVID symptoms was published (https://www.santepubliquefrance.fr/). Wölfel et al. measured the daily amount of vRNA in swab samples for few patients (27). This data was used as an estimation of viral expression for all infected people and we also assume that people with symptoms would consult 2 days after the onset of the illness, and that at any moment the number of patients with strong symptoms was proportional to the total number of infected people. Accepting the above hypothesis, the convolution of two dataset (i.e. number of consulting patient and model of excretion) is an emission-proxy, proportional to the total amount of viral RNA shed in stools in a given population.

## Results

Three major wastewater treatment plants (WWTP) managing over 75 000 cubic meters per day, were sampled at the inlet of the plant from March 5^th^ to April 23th 2020. All processed samples scored positive for the presence of SARS-CoV-2 genomes as assessed by RT-qPCR on the viral E gene. All positive samples were confirmed by RT-qPCR on the viral RdRp gene (Figure 1). The COVID-19 epidemic in the same region was illustrated by various indicators such as the total number of COVID-19 cases treated in the regional hospitals, the accumulation of hospitalized patients or the daily death toll linked to COVID 19. Based on these epidemiologic statistics and on published data on virus shedding quantity and delay, an estimated indicator of the viral excretion in the region was calculated and compared to the viral load in wastewaters.

**Figure 1:**
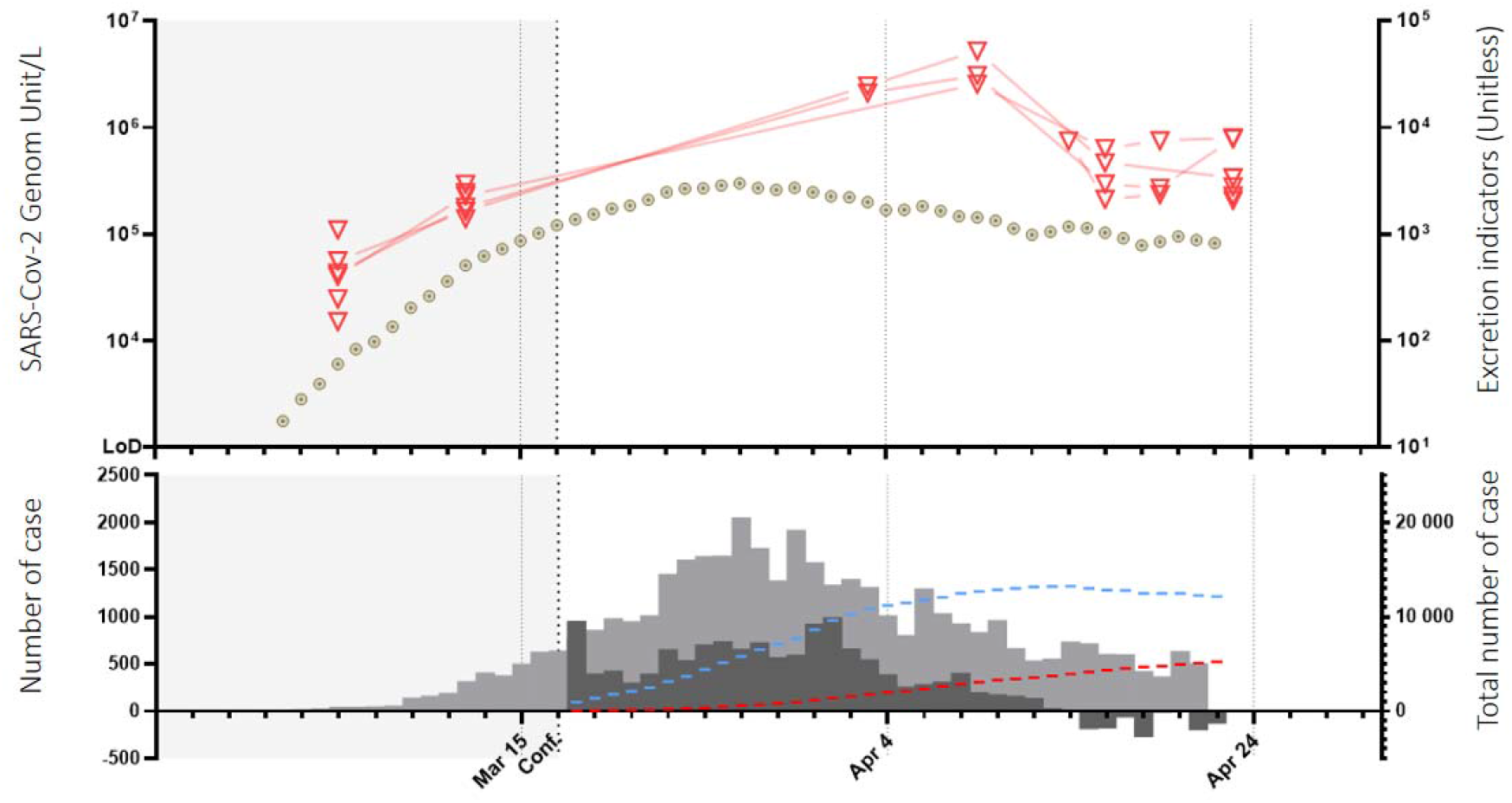
Upper panel: Quantification of SARS-Cov-2 in wastewater samples in Parisian Area in different WWTP (open inverted red triangles for important WWTP, purple open inverted triangles for smaller WWTP) in circle estimators of the viral excretion. Lower panel: In Light grey area, daily number of consultation for COVID 19 Symptoms in hospital of the Parisian area. Dark grey daily growth of hospitalized patient. Blue bar total hospitalized patient in the Parisian area, red bar cumulative deaths each day, in Parisian area. Both panel: grey background, pre-lockdown period.

Briefly, the concentration of vRNA in raw wastewater was around 5.10^4^ genome units /L on the 5^th^ o f March 2020. At the same date, less than 10 COVID-19 confirmed patients were reported and only 404 individuals were tested positive in France. For the Parisian area more specifically, 91 confirmed cases were reported at that time (on a total number of more than 12 million inhabitants) and no death was recorded. Altogether this information indicated that the COVID-19 epidemic was at an early stage in the Parisian area.

The time-course monitoring of viral load in WW displayed an exponential increase (from 5.10^4^ GU/L on March 5^th^ to 3.10^6^ GU/L, a 2-log increase in average). A peak was observed on the 9^th^ of April, followed by a marked decrease (1-log reduction in average). The shape of the concentration curve was reminiscent of the disease dynamics at the regional level, with an 8-day temporal shift.

Altogether these results underline that essential information could be obtain from wastewater epidemical monitoring, such as early starting of the epidemics, evolution of the infections, and impact of the lockdown procedures.

## Discussion

It is demonstrated here for the first time that a quantitative detection of SARS-CoV-2 in wastewaters could reflect the circulation of the virus in human populations in the Parisian area, a region called Ile-de-France. Since similar results were obtained from three independent and distant WWTP around Paris with striking similarities, the time-course survey that has been done is likely to be a direct reflect of SARS-CoV-2 dynamic in Parisian inhabitants that are connected to these WWTP. It is to note that at home lockdown is effective since March 17^th^ 2020, therefore limiting daily transport. Importantly no significant rain fall was recorded in the Parisian area that could have had an impact on virus concentration in wastewaters. More surprisingly this decrease stopped after 7 days, and the virus concentration has been stable since. This plateau is intriguing, although the emission-proxy suggests that it can partly be explained by the duration of the virus shedding period, and several hypotheses can be made. First, one may suggest that many infected people are still secreting viruses in their feces whereas the virus is not present anymore in the ORL region. This hypothesis has recently been confirmed in Chinese patients with longer periods of excretion than reported by Wölfel (7). Second, lockdown has been partial since some specific workers have been allowed to pursue essential activities that were not compatible with homeworking. These people are usually not considered at risk of severe infections (i.e. more pauci-symptomatic case), but they may promote virus circulation at a low level notably in their family if they do not strictly respect hands cleaning and mask wearing. Third, one may suggest that lockdown is not respected by few people that maintain virus circulation at a low but significant level. Virus survey in the same WWTP will likely provide some answers in the following weeks.

The observed delay between epidemiological curves in humans compared to virus quantification in wastewaters is probably due to several parameters. This may include the effective number of infected people, the timing and temporal kinetics of viral shredding in feces and other causes that are still to be investigated. Nevertheless, our data are in very good agreement with epidemiological parameters such as the number of confirmed COVID-19 patients or our excretion model. To that respect, let us note that our study provides a strong indirect evidence for a significant reduction of virus transmission in response to lockdown. According to our results, the number of people producing SARS-CoV-2 is likely underestimated when based on individual testing, especially during a pandemic where a limited quantity of virological tests did not allow for extensive testing so far. As a comparison the quantity of human enteric virus concentration in raw wastewater is around 10^6^ per liter (17).

Epidemiological investigations that have been conducted on the Diamond Princess cruise ship suggested that less than 20% of infected people were asymptomatic (12). Most of the infected people were reported to exhibit moderate nonspecific symptoms including fever, headache, body aches, intense tiredness and/or dry cough. However infected people can produce SARS-CoV-2 for a few days before the onset of symptoms and up to several days after recovery (2,7,28). Another extensive study based on Iceland population shows that 43% of SARS-CoV-2 positive patients did not report any symptoms (29). In this context, a clear majority of infected carriers may silently contaminate sensitive people. This led us to suggest that the contamination of raw wastewaters may occur before the significative appearance of clinical cases. The evolution of SARS-CoV-2 viral load in wastewater was in good agreement with the dynamics of pandemic during the first wave of infection in urbanized area, which is also in agreement with the excretion model that is proposed here. To our knowledge this is the first report demonstrating that the quantitative monitoring of SARS-CoV-2 in raw wastewater is a time-related relevant indicator of the evolution of the health status of a population linked to a sewage network. This quantitative approach was especially useful to unrevealed the dynamics of the pandemic and follow impact of government measures such as containment.

To finish, this data, if carefully utilized, could help to describe the proportion of SARS-CoV-2 excretors during all the monitored pandemic event and allow to calculate the immunity of the population, especially at the local level.

## Conclusions

Our results strongly argue for the use of a quantitative monitoring of SARS-CoV-2 genomes in urban wastewaters. This would also argue for a long-time conservation of wastewater samples in dedicated local i.e. wastewater-bank, which would allow a retrospective investigation of pathogens circulation. Additionally, wastewaters survey may provide an alternative and possibly early tool to detect pathogens in populations when investigations in humans are difficult to conduct for logistic, ethical or economic reasons, notably in poor countries that are strongly exposed to COVID-19 epidemic.

## Data Availability

This paper contain data from our own laboratory, and public data from french health agency

## Acknowledgment

Olivier Rousselot (director of “laboratoires et de l’environnement”, SIAAP) is acknowledged for his support and critical reading of the manuscript. Vincent Rocher for management of sampling at the SIAAP Innovation department. Technical assistance from the R&D Eau de Paris Laboratory members, and Alban Robin for helpful support.

## Contribution

SW, ER performed the virus measurements; SW, LM started the project; JLA, SW provided samples; LM, VM, JMM, SW for the redaction of the manuscript; YM, RT, JLA technical and theorical discussion.

## Supplementary data

None

